# Using explainable artificial intelligence to identify linguistic biomarkers of amyloid pathology in primary progressive aphasia

**DOI:** 10.1101/2024.05.02.24306657

**Authors:** Cole Robertson, Neguine Rezaii, Daisy Hochberg, Megan Quimby, Phillip Wolff, Bradford C. Dickerson

**Author notes:** corresponding author, co-first author.

## Abstract

**Introduction:** Recent success has been achieved in Alzheimer’s disease (AD) clinical trials targeting amyloid beta (β), demonstrating a reduction in the rate of cognitive decline. However, testing methods for amyloid-β positivity are currently costly or invasive, motivating the development of accessible screening approaches to steer patients toward appropriate diagnostic tests. Here, we employ a pre-trained language model (Distil-RoBERTa) to identify amyloid-β positivity from a short, connected speech sample. We further use explainable AI (XAI) methods to extract interpretable linguistic features that can be employed in clinical practice.

**Methods:** We obtained language samples from 74 patients with primary progressive aphasia (PPA) across its three variants. Amyloid-β positivity was established through the analysis of cerebrospinal fluid, amyloid PET, or autopsy. 51% of the sample was amyloid-positive. We trained Distil-RoBERTa for 16 epochs with a batch size of 6 and a learning rate of 5e−5, and used the LIME algorithm to train interpretation models to interpret the trained classifier’s inference conditions.

**Results:** Over ten runs of 10-fold cross-validation, the classifier achieved a mean accuracy of 92%, SD = 0.01. Interpretation models were able to capture the classifier’s behavior well, achieving an accuracy of 97% against classifier predictions, and uncovering several novel speech patterns that may characterize amyloid-β positivity.

**Discussion:** Our work improves previous research which indicates connected speech is a useful diagnostic input for prediction of the presence of amyloid-β in patients with PPA. Further, we leverage XAI techniques to reveal novel linguistic features that can be tested in clinical practice in the appropriate subspecialty setting. Computational linguistic analysis of connected speech shows great promise as a novel assessment method in patients with AD and related disorders.

## Introduction

Recently reported clinical trials investigating disease-modifying therapies targeting amyloid beta (Aβ) in patients with early-stage Alzheimer’s disease (AD) have shown promising evidence of clinical efficacy.^1,2^ These agents significantly reduce cerebral amyloid burden and slow clinical decline by 25-35% in 18 months. To qualify for treatment, the amyloid status of patients with MCI or mild dementia suspected of being due to AD needs to be determined. While lumbar puncture for cerebrospinal fluid analysis and amyloid PET imaging are currently the gold-standard methods for determining amyloid status *in vivo*, the invasive nature or high cost of these approaches may prevent some patients from obtaining this diagnostic information in a timely fashion. In the face of this diagnostic challenge, behavioral measures that accurately determine the likelihood of amyloid positivity can serve as a screening measure, steering patients toward the additional testing most appropriate for them. Among a large family of digital biomarkers of various types of behaviors, computational linguistic measures that can be applied to connected speech samples show promise for various purposes, including as a low-cost, accessible approach for triaging patients for undertaking further assessments that require specialty expertise or invasive or expensive biomarkers.

In this work, we aim to use machine learning (ML) methods to predict cerebral amyloid-β status (positive or negative) in patients with Primary Progressive Aphasia (PPA) using short, connected speech samples. PPA is a clinical syndrome characterized by the gradual loss of language abilities, arising when AD or Frontotemporal Lobar Degeneration (FTLD) (or rarely other neurodegenerative diseases) initially affects brain language networks.^3,4^ When PPA is the initial presentation of AD, the characteristics of a patient’s aphasia usually involve impairments in word retrieval and sentence repetition (often as a result of a deficit in auditory-verbal working memory) and may be consistent with the logopenic variant (lvPPA).^5–7^ When PPA is the initial presentation of FTLD, the patient’s aphasia is usually characterized by either impaired semantic memory (semantic variant, svPPA) or by simplified syntactic structure and effortful speech (nonfluent variant, nfvPPA).^5^ Previous work on predicting amyloid status in PPA using connected speech has reached an accuracy of 77%.^8^ This study used a theory-driven approach using a large set of lexical, syntactic, semantic, and pragmatic features for prediction. The language features that contributed to predicting a positive amyloid status included lower idea density, difficulty producing concrete nouns, and higher markers of uncertainty.

We seek to improve this state-of-the-art by building a text classifier to predict amyloid status using a pre-trained language model. We use a version of RoBERTa, a language model based on BERT^9^ but with optimized training procedures.^10^ RoBERTa was pre-trained on massive text data (over 160 GB, or 8.5–34 billion words depending on the encoding) using masked language modeling objectives (i.e., models are trained to predict withheld words using surrounding context).^10^ Through such training processes, language models can acquire in-depth knowledge of a language that can be transferred to various downstream language tasks, including the present text classification problem. In general, text classification involves training models to discriminate between classes (amyloid-β status in the present case) assigned to input data (connected speech samples).^9,11,12^ By leveraging this “transfer” of language knowledge from pre-training to downstream text classification, we hope to improve on the state of the art, which has instead relied on “feature engineering”.^13–15^ Feature engineering is the process of transforming unstructured multidimensional data (e.g., images, connected speech samples) into sets of numerical characteristics (e.g., counts of lexical, syntactic, or semantic features). These are then passed to a downstream learning algorithm. However, feature engineering can incorporate researchers’ preconceived notions about which features are pertinent to a classification task and inevitably reduce high-dimensional linguistic representations. Our approach is to improve reported text classification accuracy by training distil-RoBERTa to map directly from connected speech to amyloid-β status.

Once the task of classification is complete, we use Explainable AI (XAI) methods to identify the linguistic features that distil-RoBERTa has found to be most critical. Explainable AI research focuses on making ML algorithms interpretable so that the bases for model outputs can be understood by humans.^15,16^ Our work joins a growing literature that implements XAI methods in medical research, where professionals need to understand and trust ML model outputs before incorporating them into clinical decision-making processes.^16–22^ In this context, XAI methods can be put to various uses, including helping medical professionals choose between competing ML algorithms,^23^ uncovering and correcting model biases,^19^ or improving model performance.^24^ Our work answers calls in the literature to pursue a different approach: We use XAI for feature *discovery*.^25,26^ We first train distil-RoBERTa to classify amyloid-β status. We then use an interpretation method^27^ to discover the features (words and classes of words) that contribute most to our classifier’s decisions. Finally, we use dimension reduction and data visualization techniques to “see” the critical determinants of the classifier’s inference process.

Using XAI is advantageous for two reasons. First, it achieves a balance between performance and transparency. As ML models grow more complex, they often achieve better performance at the cost of interpretability, i.e., the best explanation of a simple model is the model itself. For example, a coefficient (and intercept) in a univariate linear regression captures everything the model “knows” about the relationship between a predictor and an outcome. Increasing model complexity can confer gains in performance, but models tend to become less inherently explainable. Pretrained deep language models (like RoBERTa) are paradigmatic examples of highly complex but inherently unexplainable models, often comprising millions or billions of unintelligible parameters. By leveraging XAI methods, we retain performance gains due to increased complexity without sacrificing interpretability.^27–29^ Machine learning models are good at identifying patterns in data that humans cannot comprehend, but if models are uninterpretable, this knowledge remains locked away in millions of incomprehensible parameters. We employ XAI techniques to cut this Gordian Knot by allowing us to “discover” the speech patterns RoBERTa has identified as characterizing amyloid-positivity without being limited to a small set of pre-engineered features.

## 1 Methods

### 1.1 Participants

For this cross-sectional study, we analyzed data from 71 patients with PPA from our ongoing longitudinal study in the PPA Program of the Frontotemporal Disorders Unit at Massachusetts General Hospital (MGH). All patients received a standard clinical evaluation comprising a structured history obtained from both patient and informant, comprehensive medical, neurological, and psychiatric history and exams, neuropsychological and speech-language assessments, and a clinical brain MRI that was visually inspected for 1) regional atrophy consistent with or not consistent with a given syndromic diagnosis, and 2) other focal brain lesions or evidence of cerebrovascular disease. The clinical formulation was performed through consensus conference by our multidisciplinary team of neurologists, psychiatrists, neuropsychologists, and speech-language pathologists, with each patient classified based on all available clinical information as having mild cognitive impairment or dementia (Cognitive Functional Status), Cognitive-Behavioral Syndrome, and likely etiologic diagnosis ^30^. All patients included in this study met the diagnostic criteria for PPA and were subtyped, if possible, into one of the major subtypes with a clinical imaging-supported atrophy pattern: the nonfluent/agrammatic variant (nfvPPA), semantic variant (svPPA), or logopenic variant (lvPPA) of PPA ^5^. Furthermore, all patients also met the following criteria: (1) no focal brain lesions or significant cerebrovascular disease (e.g., previous strokes, cerebral hemorrhages, meningiomas); (2) no major psychiatric illness not adequately treated; and (3) native speaker of English. The clinical and demographic information on the sample of participants is shown in Table 1, which included nfvPPA (n = 18), svPPA (n = 18), lvPPA (n = 32), and mixed PPA (n = 3). Ratings on the Progressive Aphasia Severity Scale (PASS) were also included to illustrate the severity of the patient’s aphasia at a more fine-grained level than the Clinical Dementia Rating supplemental language box ^31^. The PASS uses the clinician’s best judgment and integrates information from the patient’s test performance and a structured interview with the patient and an informant. The PASS includes “boxes” for fluency, syntax, word retrieval and expression, repetition, auditory comprehension, single-word comprehension, reading, writing, and functional communication. The PASS Sum-of-Boxes (SoB) is the sum of the box scores. The clinical and demographic information on the patients is shown in Table 1. There were no significant differences between the PPA-AD and PPA-FTLD groups in the variables included in Table 1. All study participants provided informed consent in accordance with guidelines established by the Mass General Brigham Healthcare System Institutional Review Boards, which govern human subjects research at MGH and specifically approved this entire study.

**Table 1.** The clinical and demographic information of patients with amyloid positive and negative status

| Amyloid status | Amyloid positive | Amyloid negative |
| --- | --- | --- |
| Age of symptom onset (SD) | 70.5 (8.5) | 66.9 (6.1) |
| Sex, percent female | 44.4% | 51.4% |
| Years of education (SD) | 16.1 (2.5) | 16.5 (2.6) |
| Handedness, percent right-handed | 86.2% | 84.4% |
| PASS, Sum of Boxes (SD) | 5.9 (3.4) | 4.2 (2.9) |

#### Determining amyloid status

We determined amyloid status by examining cerebrospinal fluid (CSF) analysis, amyloid PET scan and/or autopsy results. In this cohort, 36 patients (51%) were amyloid positive and 35 individuals were amyloid negative (49%).

For patients who underwent CSF biomarker analysis, CSF was obtained through lumbar puncture and was sent to Athena Laboratories for ADMARK analysis for levels of amyloid-β, total tau, and hyperphosphorylated tau. Standard reference cutpoints were used to determine whether the levels were abnormal in a manner consistent with AD neuropathologic changes.

Some participants underwent ^11^C-Pittsburgh Compound B (PiB) PET scans. The PiB radiotracer was prepared as described previously.^32^ All PET data were acquired using a Siemens (Knoxville, TN) ECAT HR+ scanner: 3D mode, 63 imaging planes, 15.2 cm axial field of view, 5.6 mm transaxial resolution, and .4 mm slice interval. PiB PET images were acquired with an 8.5 to 10.5 mCi bolus injection followed immediately by a 40-60 min acquisition. All PET data were reconstructed and attenuation corrected; each frame was evaluated to verify adequate count statistics and interframe head motion was corrected.

Amyloid-β positivity was determined by visual read according to previously published procedures ^33^ as well as a summary distribution volume ratio (DVR) of frontal, lateral temporoparietal, and retrosplenial (FLR) regions greater than 1.2.^34^

Autopsies were performed according to standardized protocols at the Massachusetts Alzheimer’s Disease Research Center.^35^ At the time of autopsy, brains were divided at the midline, with one hemisphere sectioned and frozen at −80°C and the other fixed in 10% buffered formalin. After 10 to 14 days, the formalin-fixed hemisphere was sectioned, photographed, and evaluated grossly by a board-certified neuropathologist. In each case, 25 tissue sections were obtained utilizing a blocking protocol that widely samples anatomical regions relevant for diagnosis of neurodegenerative disorders. These regions included the hippocampus, thalamus, subthalamic nucleus, basal ganglia, amygdala, cerebellum with dentate nucleus, and all levels of the brainstem. Multiple sections of frontal, parietal, temporal, cingulate, and calcarine cortices were also sampled. The tissue blocks were processed on a Thermo Scientific Excelsior ES tissue processor, and embedded in paraffin. All sections were cut on a microtome at 7 μmol/l and stained with Luxol fast blue/haematoxylin and eosin for routine assessment. Bielschowsky silver stain was carried out on sections from select blocks. Immunohistochemistry was performed on sections from select blocks and processed on a Leica Bond RX automated stainer (Leica Biosystems), with anti-human pan-tau antibody (Dako), anti-human beta-amyloid antibody (Dako), anti-GFAP antibody (Sigma-Aldrich), anti-TDP-43 antibody (Proteintech), and anti-synuclein antibody (Thermo Scientific), as well as anti-ubiquitin antibody in the case of Subject 11 (ThermoFisher). Neuropathological examination was performed in accordance with published guidelines.^36,37^

### 1.2 Classification

Since our task is a small-data classification problem, we use a “distilled” version of RoBERTa (“distil-RoBERTa”). Distillation involves training a smaller model to learn representations in a larger model to achieve comparable results but with less computational cost.^13^ This smaller model makes sense to avoid over-fitting.^14^ We connected distil-RoBERTa to a standard fully connected classification layer and trained on amyloid status labels (negative vs. positive) using *ktrain* version 0.33.0^38^ in Python 3.7.15. We trained for 16 epochs with a batch size of 6 and a learning rate of 5e−5. To assess generalizability, we performed ten runs of 10-fold cross-validation. To avoid over-fitting on the training data, we rolled back model weights to the best epoch by validation accuracy. If multiple epochs had equal validation accuracy, we chose the epoch with the lowest validation loss. The interpretation algorithm necessitated we choose the best run, which we did accordingly: *R_jk_* = *argmax*(R*^n^* ^=10^), which is to say we calculated the best *n* = 10 training runs for each *j* = 4 classification metric (accuracy, precision, recall, F1) and *k* = 2 class label (positive, negative). To get the best run, we took the mode of the resultant eight metrics (8/8 agreed).

### 1.3 Interpretation

#### 1.3.1 LIME

To establish which linguistic features determined the classifier’s inferences, we used Local Interpretable Model-agnostic Explanations (LIME)^27^ implemented in the Python package *eli5,* version 0.13.0.^39^ The LIME algorithm works by training an interpretable “white box” classifier to predict the probabilities output by an uninterpretable “black box” classifier (the best run of the trained distil-RoBERTA-based classifier, in this case). The notion is that complex non-linear classification functions can be *locally* approximated in the region of a specific example by a linear model (see Figure 3 in Ribeiro et al.^27^). In order to retain interpretability, LIME uses binary vectors to indicate the presence of absence of a lexical item, i.e. *x*^′^ ∈ {0,1}*^d^*^′27^ (i.e. in place of word embeddings, *x* ∈ R*^d^*, which are incomprehensible).

**Figure 3:**
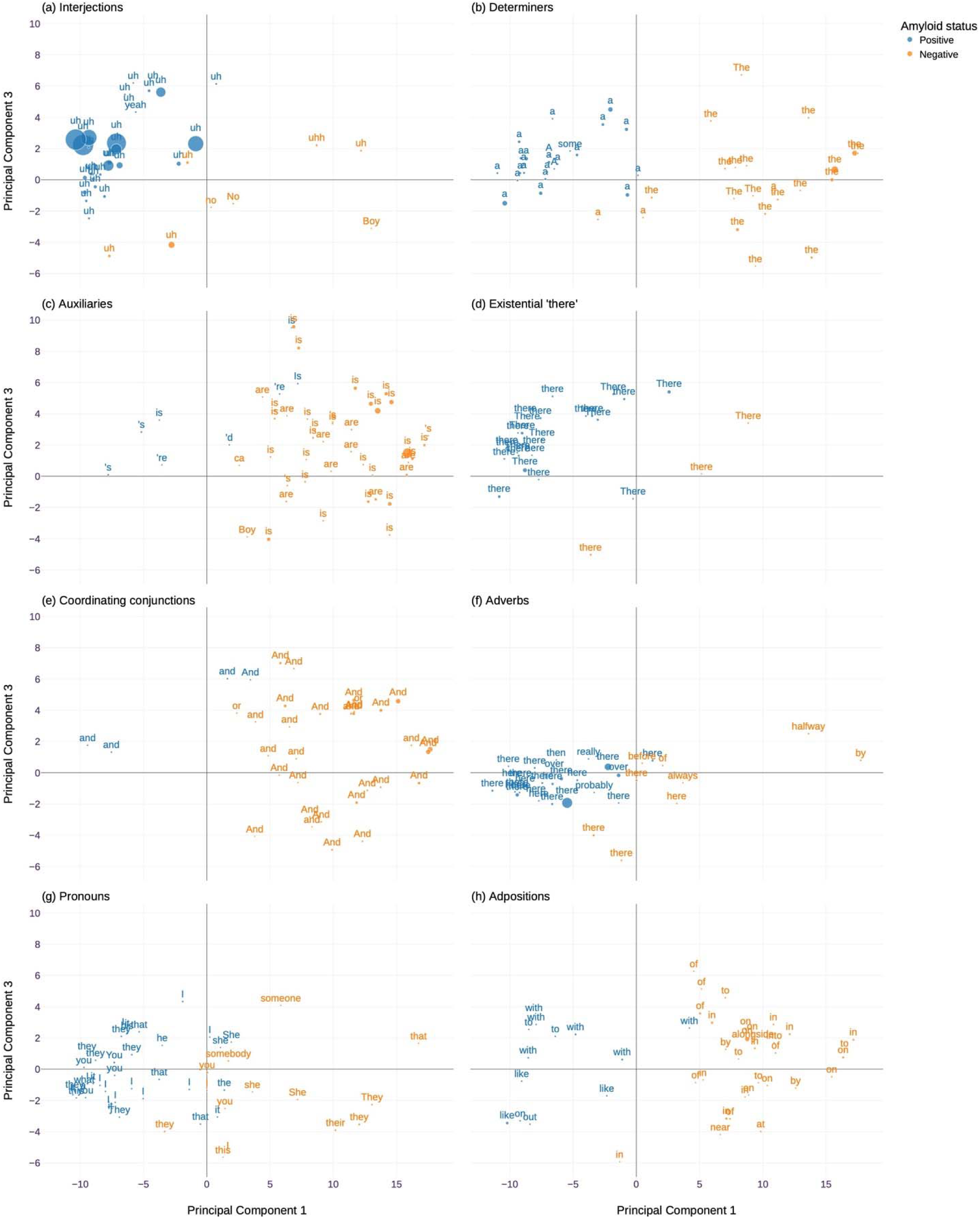
*The top* n *= 5 words within each tag for the top 8 POS tags (apart from verbs and nouns) in descending order of* S’ *from Equation 2*.

Each text input is then systematically permuted (different words are removed). Simple word counts are passed to a linear model (a logistic regression) trained to predict the resultant changes in the black-box classifier’s output probabilities. This allows LIME to derive a “weight” for each lexical item which captures how much impact it has on the black-box classifier’s probabilities (inference conditions). It may be helpful to conceive of weights as regression coefficients which capture change in *y* (classifier output probabilities) as a function of change in *X* (the presence or absence of lexical items in input texts).^27^

#### 1.3.2 Feature discovery

Critically, LIME explanations are *local*, which presents a data-visualization challenge: How can global behavior be represented when, even for small data sets, humans cannot be expected to exhaustively inspect all *N* local explanations? We addressed this challenge in two ways. Firstly, we implemented the “pick” algorithm proposed in Ribeiro et al.,^27^ which chooses the *B* most informative local explanations, where *B* is the number of explanations humans can reasonably be expected to inspect, here *B* = 2 (for each class).

Second, we used dimension reduction, combined with techniques from Natural Language Processing (NLP), to derive the lexemes *and* Parts of Speech (POS) which most impact the classifier’s inferences. A part of speech, or word class, is the grammatical role a given lexical item plays in a particular sentence, e.g., noun, verb, determiner, etc. We used the *en core web lg* model, version 3.4.1, in *spaCy*, version 3.4.3, to assign a part-of-speech tag to each word in the input texts. Specifically, we used the *token.pos_* attribute in *spaCy*, which uses the Universal Dependencies POS taxonomy.^40^ We modified this slightly to add an “existential there” category, “EX”. This allowed for differentiation between existential uses of *there*, e.g., *There is a tree* and adverbial ones, e.g., *He’s over there by the boat*. This was done because an inspection of the data indicated existential there was an important category in our data which was not captured in the universal dependencies.

We then defined a function *f*(*t*) which ingests text, *t*, discards punctuation (since text examples were transcribed from audio recordings, punctuation may have represented transcribers’ idiosyncratic practices), and concatenates the *spaCy* POS tag with each word, e.g., *f*(*Over there, there is a tree.*) = *Over*_adp_ *there*_adv_ *there*_ex_ *is*_verb_ *a*_det_ *tree*_noun_. Passing the output of *f*(*t*) to LIME allowed the effect of each unique word-POS pair on the classifier’s output probabilities to be estimated independently, e.g., separate weights would be derived for *there*_ex_ and *there*_adv_ in the example above.

To visualize these words in the semantic space of the classifier, we extracted the embeddings from the second-to-last layer (which is connected to the classifier head) of the trained classifier for each fold. To align these with word-POS pairs, some averaging was necessary. This was because distil-RoBERTa uses sub-word tokenization: Words which are out of vocabulary are broken down into constituent parts and embedded, e.g., *symbolism* might be represented with an embedding for each of *symbol* and *ism*. These needed to be aligned to the *spaCy* tokens, which are words. To do this, we calculated the mean of all subword tokens within the span defined by the *spaCy* token boundaries, i.e. for each token, *j*, an embedding was calculated by 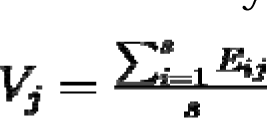, where *s* is the number of subword tokens in the span of each word, and *E_ij_* is the context-sensitive embedding of the *i*_th_ subword token of the *j*_th_ word. This resulted in each text being represented as an *m* by *n*, matrix, *T* where *m* = *dim*(*f*(*t*)), and *n* = 768, the distil-RoBERTa embedding dimensions, R^768^.

We matched each matrix *T_i_* for each *i* participant with the LIME weights (this was possible since *f*(*t*) was used both to extract embeddings and calculate LIME weights). Some averaging was needed to facilitate this: Some token weights are 0, and LIME only derives one weight for each orthographically unique token in *f*(*t*), e.g. there would only be one weight for “ball” in “The boy with the ball throws the ball to the girl”, whereas there would be 2 (slightly) different context-sensitive embeddings. We therefore defined a set *S_i_* = ∃{*f*(*t_i_*)} : *W_ij_ >* 0 which captured the set of *j* unique token types with non-zero weights *W_ij_* in *f*(*t*) for each *i* participant. We calculated the mean of the token embeddings across these, 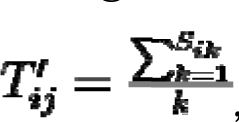, where *k* is the number of tokens in *f*(*t_i_*) which are orthographically identical to each *j* token type where *W_ij_ >* 0 in *S_i_*.

We concatenated the *T^‣^_i_* matrices for each *i* participants into an embedding matrix *M_Si_N,*_R_768. As a visualization convenience, we used singular value decomposition to reduce the second dimension of the *M* to R^3^. Inspection of the scree-plot indicated most variance was explained by the first factor, which appeared to relate to the class labels, see Supplementary Materials (SM). This resulted in *N* = 3673 observations of 816 unique linguistic features (word-POS pairs) with non-zero weights.

## 2 Results

### 2.1 Classification

The classifier was able to differentiate between amyloid positive and negative patients with a mean accuracy score of 0.92, *SD* = 0.02 (see Table 2). This was significantly better than chance, χ^2^(1*, N* = 71) = 55.94*, p <* 0.001. An accuracy of 92% represents an absolute improvement of 15% over previously published accuracy identifying amyloidosis from connected speech, i.e., 77% in Slegers et al.^8^ Additionally, class labels in the latter work were imbalanced: 40.17% of patients were amyloid-positive, compared with 51% in this study. Using the zero-rule baseline classifier (which always guesses the most frequent class label), our results improve over baseline by 42.1%, compared with 17.17% in Slegers et al.,^8^ a relative improvement of 145.16%.

**Table 2.** Distil-RoBERTa classification metrics

| Amyloid status | Accuracy* | Precision** | Recall<sup>†</sup> | F1<sup>‡</sup> |
| --- | --- | --- | --- | --- |
| Negative | 0.92 (0.01) | 0.92 (0.03) | 0.93 (0.02) | 0.92 (0.02) |
| Positive |  | 0.93 (0.02) | 0.92 (0.02) | 0.92 (0.02) |

Mean and (SD) are presented over 10 training runs.
\*Accuracy is $a = (tp+tn)/(tp+fp+fn+tn)$ where $tp$ is the number of true positives, $tn$ is the number of true negatives, $fp$ is the number of false positives and $fn$ is the number of false negatives.
\*\*Precision is $p = tp/(tp + fp)$ ; it captures the model's likelihood of being correct if it makes a positive prediction and is therefore sensitive to the model's type I error rate.
<sup>†</sup>The denominator in the recall is false negatives, $r = tp/(tp+fn)$ ; it, therefore, captures whether the model tends to miss true examples and is sensitive to the model's type II error rate.
<sup>‡</sup> $F1$ is the harmonic mean of recall and precision, $F1 = \frac{2rp}{r+p}$ , and attempts to balance the two.

### 2.2 Interpretation

#### 2.2.1 LIME

LIME works by using simple interpretable models to linearly approximate a black-box classifier’s classification function in the local region of a given example. It is therefore important to understand how well the interpretation models capture the black box model’s behavior. We do this in two ways.

First, we calculate the performance of the interpretation models against the classifier’s output probabilities for the *n* = 5000 permuted text inputs generated by the LIME algorithm. On average, our interpretable models performed well: Mean Kullback-Leibler divergence was ^-^X = 0.03*, SD* = 0.03 (low values indicate high agreement between two distributions). Mean distance-weighted-accuracy was ^-^X = 0*.95, SD* = 0.05. This is accuracy weighted by a distance measure (cosine similarity), and penalizes permuted text examples which are distant from the true example.^27^ Both metrics indicate that the interpretable models do a good job, on average, of capturing the classifier’s behavior.

Second, we report classification metrics for the interpretation models’ predictions on the actual text examples, against both the classifier’s predicted labels and ground truth (Table 3). Again, these suggest the interpretable models do a good job of capturing the classifier’s predictions and the true labels.

**Table 3.**
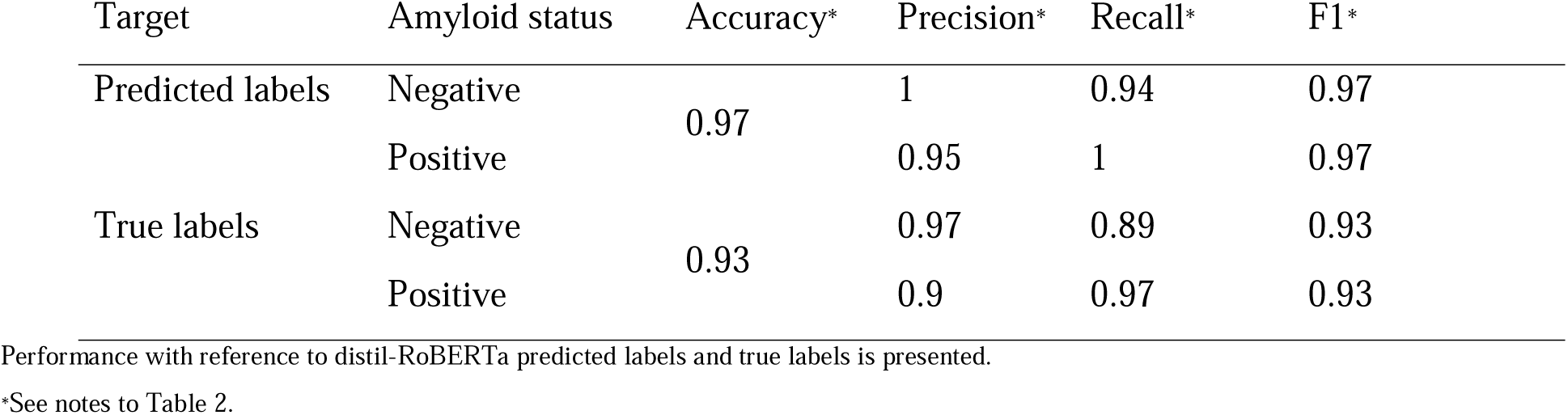
Interpretable LIME models’ classification metrics

#### 2.2.2 Feature importance by part of speech

We first wanted to understand which parts of speech in general weighted most highly on our classifier’s inference conditions, i.e. which parts of speech were the most important basis for its predictions. Selecting the top POS tags was non-trivial since frequency differences between different parts of speech in English affects results. For instance, the large number of orthographically unique nouns and verbs were associated with a large number of unique but small weights, whereas the small number of orthographically-unique, but inferentially important, interjections were associated with a small number of unique but large weights. Summing the weights by POS tag would over-represent the importance of nouns and under-represent the importance of interjections while aggregating using the mean would skew results in the other direction. We balanced these concerns by ranking POS tags using a score:

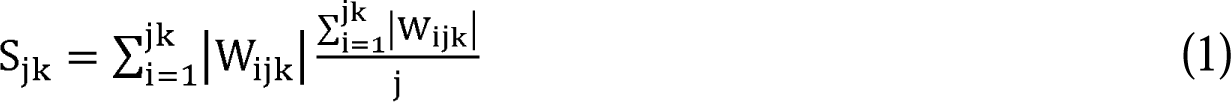

for each POS tag, *j,* observation, *i*, and class label, k, i.e. the product of the sum and the mean of the absolute value of the weights, for each POS tag and label. Because it is a product of the absolute value of the weights, *S* is not directional; it simply attempts to rank POS tags in terms of their importance to classifier inference conditions for each label. Results suggest the 5 most critical POS tags for amyloid positive individuals are interjections, determiners, verbs, nouns, and existential ‘there’. For amyloid negativity, it is determiners, nouns, interjections, auxiliaries, and verbs (Fig. 1). This suggests clinicians seeking to diagnose amyloid positivity should pay close attention to these parts of speech. In the next section, we look more granularily at language production patterns characteristic of amyloid status.

**Figure 1:**
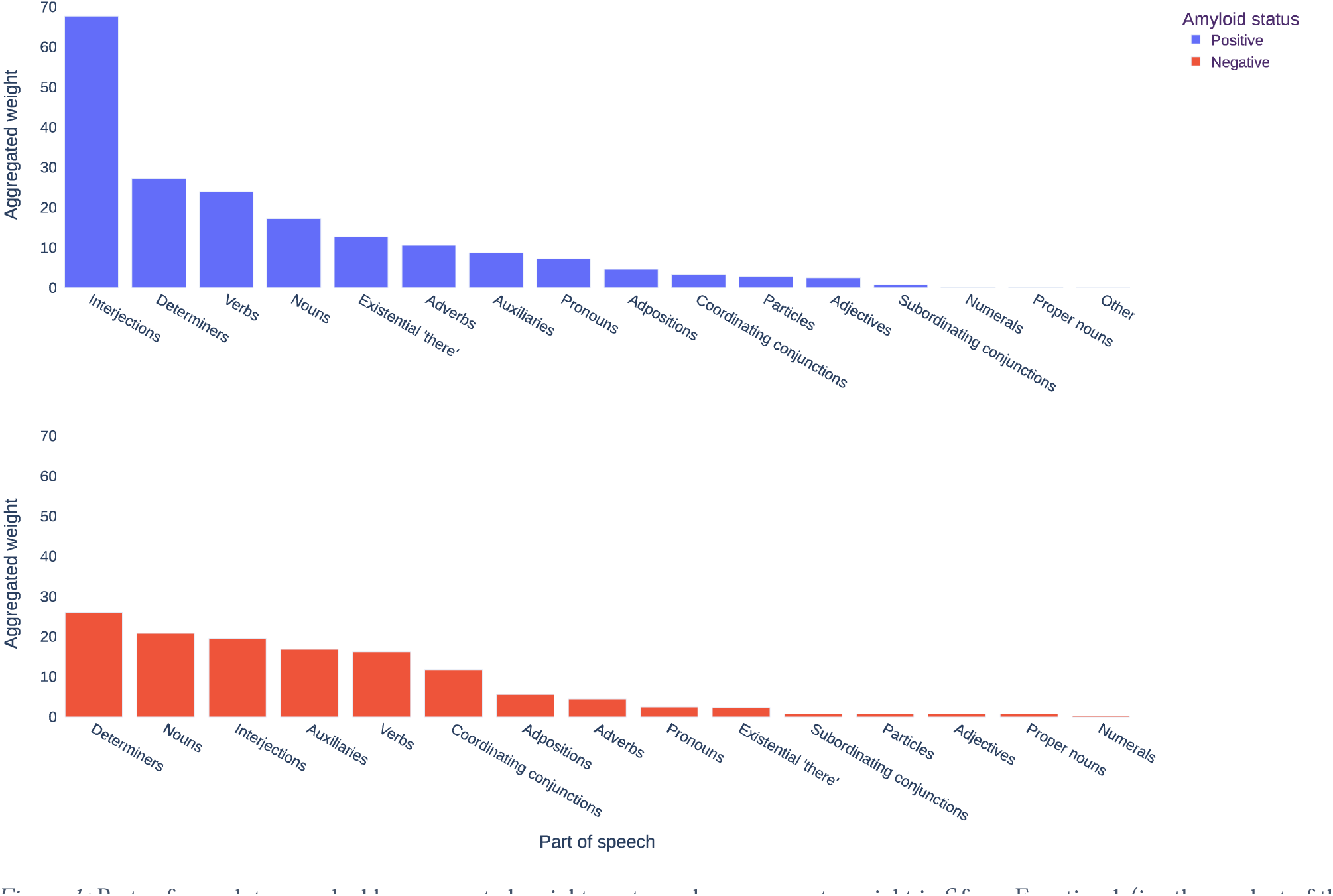
Parts of speech tags ranked by aggregated weight per tag, where aggregate weight is *S* from Equation 1 (i.e. the product of the sum and the mean of weight per tag).

#### 2.2.3 Feature discovery

We deemed the utility of presenting orthographic representations of tokens to be high, i.e. we wanted to plot the important words. To do so, intelligibility necessitated we did not plot all the words. Therefore, we used the weights to select the top *n* tokens by LIME weight within each participant’s connected speech sample. For verbs and nouns, *n=*2, because they are high-frequency, Fig. 2. For other POS tags, *n*=5, Fig. 3. Not all POS tags are plotted (SM), rather, after verbs and nouns, we plot the top 8 tags across both class labels by *S* from Equation 1, Fig. 3. Crucially, Fig. 2 & 3 *sometimes* indicate a difference in the frequency with which amyloid positive/negative patients use a given POS, e.g., amyloid positive patients use more injections (Fig. 3a). In *other* cases, a difference of usage within POS tag is indicated, e.g., amyloid positive patients use more indefinite articles “a” and fewer definite articles “the” (Fig. 3b). To give examples of the features in context, Fig. 4 presents the top explanations for each class label using the submodular pick algorithm from Ribeiro et al.,^27^ which is designed to yield the set of examples with the best coverage and least redundancy against the full dataset. Finally, we also release an interactive visualization dashboard of these figures, available at <<TKTKTKT>>. We encourage readers to use it, as it provides interactive features which are not possible offline.

**Figure 2:**
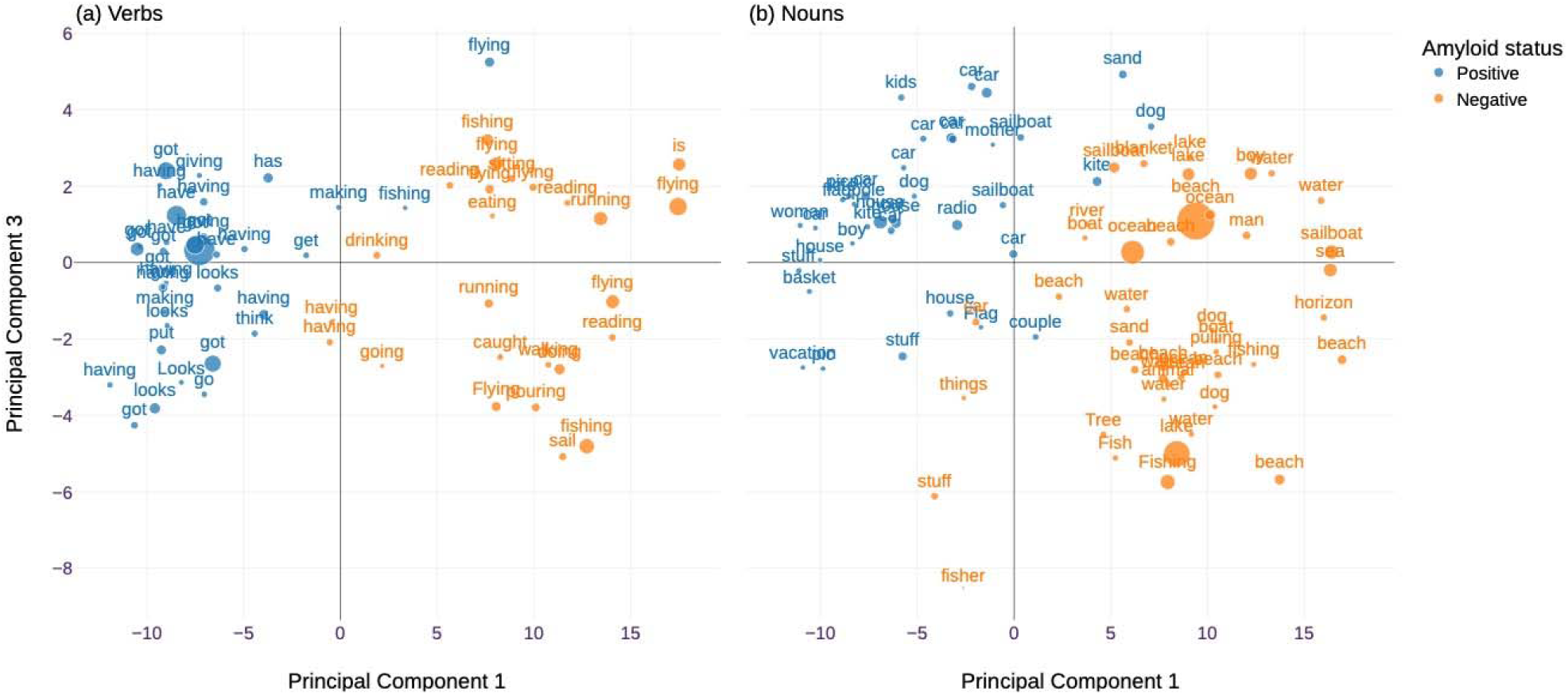
The (a) verbs and (b) nouns from among the top 2 words by LIME weight in each participant’s connected speech sample. Above and Figs. 3 & 4, we plot tokens over principal components 1 and 3 (for aesthetics reasons, SM). We use point size to represent LIME weights using the following monotonic transformation: ^r^ <u>^Wl-min(W)^</u>, where λ is a constant scaling factor chosen for aesthetic max(W)-min(W) reasons to equal 5, and W is the vector of LIME weights.

**Figure 4:**
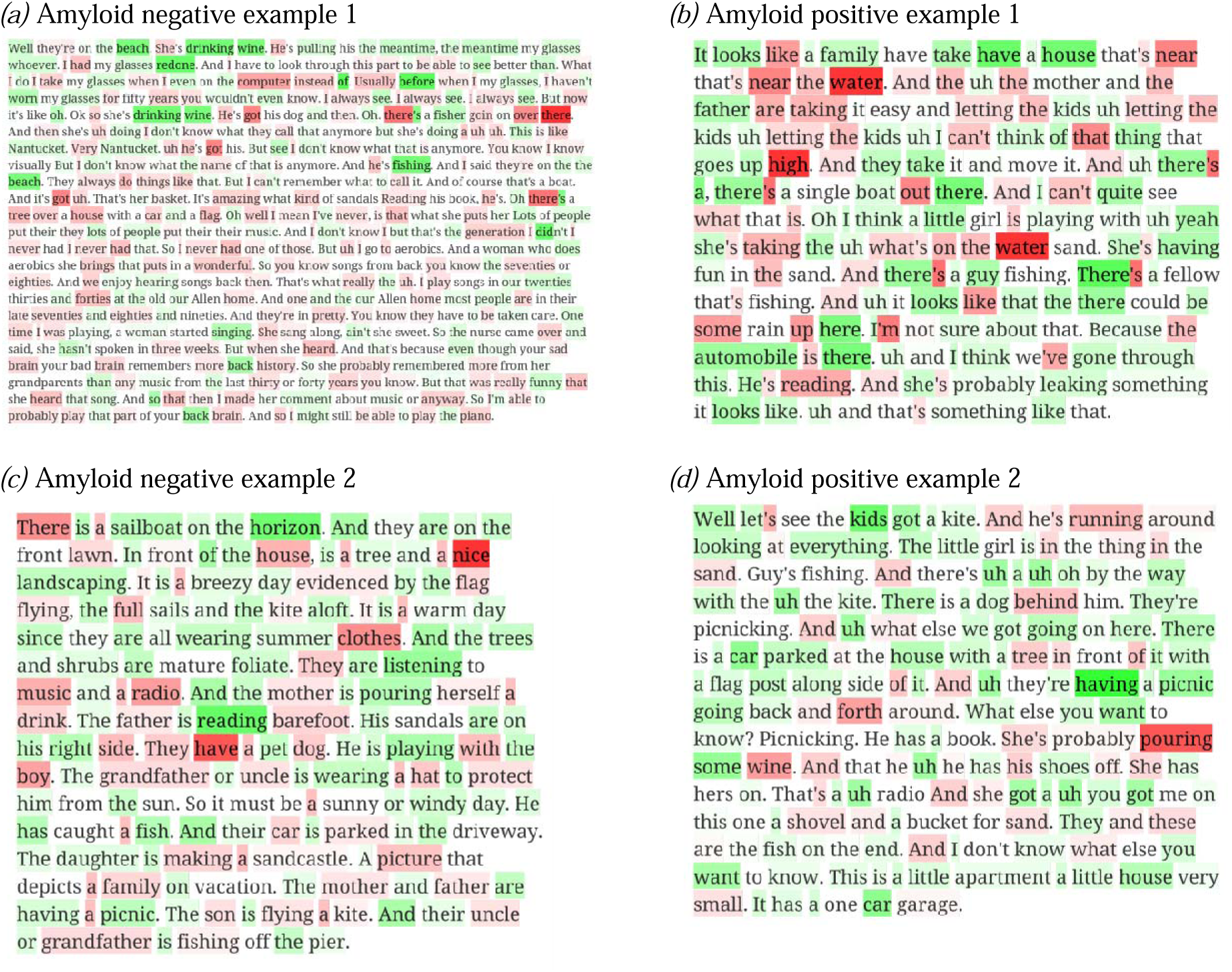
The top 2 most informative examples for each label, according to the submodular pick algorithm from Ribeiro et al. 2016.^27^ These examples were chosen from the subset of examples where y = ŷ = y^′^, i.e. where true labels equal classifier predictions equal interpretation-model predictions. Green highlights indicate a word that positively increments the probability of the predicted clas, i.e. in amyloid positive examples the inclusion of green-highlighted words increases the classifier’s predicted probability that y = amyloid positive. In negative examples, the opposite is true.

## 3 Discussion

This study aimed to use explainable artificial intelligence to predict amyloid status in PPA patients based on their short, connected speech samples and to identify linguistic biomarkers of amyloid pathology. Our approach showed significant improvements in several key areas when compared to prior research on this topic. Using distil-RoBERTa for the classification task, we achieved an accuracy of 92%. This result surpasses the accuracy reported in a previous study on predicting amyloid status using connected speech samples, exhibiting a relative improvement of 145% when adjusting the chance level associated with each study.^8^ Furthermore, leveraging a pre-trained language model for classification eliminated the need to rely on a set of engineered features. Rather, classification was carried out using the model’s massive prior knowledge of language, which was fine-tuned to suit our specific classification task. After accomplishing the task, we interrogated the model using XAI to uncover the features that discriminated between amyloid positive and negative speech samples, allowing for the discovery of features previously unnoticed by researchers and clinicians. This approach is of particular importance given the paucity of language features distinctive of the amyloid-positive subtype of PPA, mainly lvPPA.^13^ It has also been shown that the connected speech of patients with lvPPA has similarities in several key syntactic and lexical domains with patients with svPPA^41^ which emphasizes the need to discover linguistic features that are more specific to amyloid-positive status. In what follows, we discuss some of the features made explicit using this approach.

One notable difference between amyloid-positive and amyloid-negative patients with PPA is their use of verbs. Specifically, amyloid-positive patients tended to use abstract or semantically underspecified verbs such as “get,” “have,” and “make,” while amyloid-negative patients used more concrete verbs like “pour,” “sail,” and “fly.” This distinction could be interpreted in several ways. One possible explanation is that of light versus heavy verb construction.^42^ Some abstract verbs, such as “have,” “go,” and “make,” can be used in a light verb construction (LVC) where a noun or adjective accompanies them to create a new meaning. Examples of LVC include “take a walk” or “have a nap.” However, upon inspection of the abstract verbs used by amyloid-positive patients, we found a mixed pattern of LVC and non-LVC, making this construction a less likely distinguishing explanation. As shown in examples from Figure 3, the verb “have” was used both in an LVC (e.g., “having a picnic”) and, also in a non-LVC (e.g., “having a noisy radio”). Another possibility is that the distinction between the types of verbs used by each group is related to their frequency of use. Abstract verbs are among the most frequent verbs in most languages.^43^ Therefore, amyloid-positive patients may be using more high-frequency verbs than amyloid-negative patients. This interpretation is consistent with the finding by Slegers et al., 2021 who reported verb frequency as a distinctive feature between amyloid positive and negative classes.^8^ In our recent work, we also showed that verb frequency is a variable that significantly increases classification accuracy among the three variants of PPA ^44^.

Additionally, we observed conceptual differences in the category of nouns used by the two classes. Amyloid-positive patients tended to use nouns such as “radio,” “kite,” “car,” and “boy,” as well as more general terms like “stuff” and “thing.” In contrast, these patients were less likely to use words such as “water,” “beach,” and “fisher.” This distinction could potentially be explained by noun frequency, as it appears that amyloid-positive patients tend to use nouns that, on average, have a higher frequency of occurrence than other groups. However, prior research has shown that high-frequency nouns are a common feature of patients with both lvPPA and svPPA,^45^ making noun frequency a less distinctive feature. One possible explanation for this noun distinction is that patients with amyloid positivity may struggle with visuospatial processing, making them vulnerable to disregarding contextual information. The WAB Picnic Picture is a complex scene featuring multiple events taking place across different spatial layers. In the foreground, a couple is seen sitting on a picnic blanket next to a radio, with a boy flying a kite immediately behind them and a car parked in the near background. In the farthest distance, there is a beach scene with a girl building a sandcastle, a man fishing, and a sailboat on the lake. From the nouns distinctive of the two classes, it was evident that amyloid-positive patients tended to focus more on foreground elements while paying less attention to background details. This finding aligns with a meta-analysis indicating that individuals with lvPPA typically have poorer visuospatial functioning and attention than those with svPPA and nfvPPA.^46,47^

Furthermore, our bottom-up analysis of language found filled pauses such as “uh” to be predominantly used by amyloid-positive patients, consistent with similar reports in patients with lvPPA.^45,48,49^ The analysis of determiners, as another contributor to the classification, revealed an interesting pattern. Amyloid-positive patients predominantly used the indefinite article “a” over the definite article “the,” while the opposite pattern was seen in the amyloid-negative group. There are various ways to interpret this finding. One possible explanation is based on the Accessibility Theory, ^50^ which suggests that the choice of an article depends on the accessibility of the noun phrase in memory. According to this theory, the definite article is used when the noun phrase is highly accessible, while the indefinite article is used when the noun phrase is less accessible. Given the core deficit of word retrieval in patients with lvPPA, it is reasonable to expect that they tend to use indefinite articles over definite articles. Another possible explanation is based on the Given-New Contract Theory, which proposes that speakers and listeners have a shared understanding of the given and new information in a conversation.^51^ According to this theory, the use of the definite article signals that the noun phrase refers to a given or previously mentioned entity, while the indefinite article signals that the noun phrase refers to a new or unknown entity. It is possible that the differences in article usage reflect impairments in the theory of mind, which is altered in the early stages of Alzheimer’s dementia.^52^

The discussion above highlights that, by using XAI, the classification of language based on the underlying pathology is not merely a practical problem that is solved through a black-box AI model. Rather, this approach provides insights into the inner workings of the model and the potential cognitive processes underlying language features distinctive of amyloid pathology. In addition, the broad accessibility and low cost of this tool mean it has the potential to be used as a screening tool for individuals suspected of having Alzheimer’s pathology and who may be candidates for treatments with disease-modifying agents. Based on current amyloid imaging guidelines, a complete clinical examination is recommended before undergoing scanning for patients presenting with language symptoms suggestive of PPA.^53,54^ However, clinical diagnosis and cognitive testing have shown limited diagnostic performance in identifying the underlying pathology, with a sensitivity of 78% and specificity of 80%.^55^ The use of XAI has the potential to improve diagnostic accuracy and provide a more comprehensive understanding of the cognitive mechanisms underlying PPA, which could ultimately lead to better clinical outcomes for patients.

Future research is needed to overcome the limitations of this study, such as the relatively small sample size that may hinder the generalizability of the findings across diverse populations, and the complexity of the machine learning algorithms, which could result in model overfitting. Moreover, the current findings are exclusively based on English language data. To determine if the observed verb frequency effects are consistent across languages, particularly those with varying verb distributions such as Persian,^56^ future studies should include languages other than English. Lastly, additional testing in a different cohort of PPA patients is essential to validate and broaden the applicability of these findings across diverse patient groups.

## Funding

This work was supported by the US National Institute on Deafness and Other Communication Disorders grants R01 DC014296 to BCD and R21 DC019567 to BCD and PW, National Institute on Aging grants R01 AG081249 to BCD and R21 AG073744 to BCD and PW, National Institute of Neurological Disorders and Stroke grant RF1 NS131395 to BCD, and Alzheimer’s Association grant 23AACSF-1029880 and MGH Screening Technologies in Primary Care Innovation Fund (PCIF) 2023A063002 to NR. This research was carried out in part at the Athinoula A. Martinos Center for Biomedical Imaging at the MGH, using resources provided by the Center for Functional Neuroimaging Technologies, P41EB015896, a P41 Biotechnology Resource Grant supported by the National Institute of Biomedical Imaging and Bioengineering (NIBIB), National Institutes of Health. This work also involved the use of instrumentation supported by the NIH Shared Instrumentation Grant Program and/or High-End Instrumentation Grant Program, specifically, grant number(s) S10RR021110, S10RR023043, S10RR023401.

## Data Availability

Data will be accessible upon requests to Dr. Bradford Dickerson at

## Notes

### Competing Interest Statement

The authors have declared no competing interest.

### Author Declarations

All study participants provided informed consent in accordance with guidelines established by the Mass General Brigham Healthcare System Institutional Review Boards, which govern human subjects research at Mass General Hospital and specifically approved this entire study.

